# Population health impacts from the taxation of salt and sugar in the United Kingdom

**DOI:** 10.1101/2023.08.16.23294155

**Authors:** Patricia Eustachio Colombo, James Milner, Silvia Pastorino, Rosemary Green

## Abstract

**Objective:** To estimate the potential health benefits from the reduction in consumption of salt and sugar following the introduction of a proposed tax on salt and sugar in the United Kingdom (UK).

**Design:** Epidemiological modelling study. Life-table modelling was used to estimate the expected population health benefits from the reduction in consumption of salt and sugar for four scenarios, each reflecting different manufacturer and consumer responses the proposed tax. Relative risks for 24 disease-risk pairs were applied, exploring different pathways between salt and sugar consumption, and mortality and morbidity.

**Setting:** UK.

**Participants:** Population of the UK.

**Results:** The results show that life expectancy in the UK could be increased by 1.7 (0.3-3.6) to 4.9 (1.0-9.4) months, depending on the degree of industry and consumer response to the tax. The tax could also lead to up to nearly 2 (0.4-3.6) million fewer cases of preventable chronic diseases and an increase of as much as 3.5 (0.8-6.4) million years of life gained. The largest health benefits would accrue from reduced mortality and morbidity from cardiovascular diseases.

**Conclusions:** Significant benefits to population health could be expected from extending the current tax on sugar sweetened beverages to other sugary foods and from adding a tax on foods high in salt. The proposed dietary changes are likely to be insufficient to reach national public health targets; hence, additional measures to reduce the burden of chronic disease in the UK will be equally critical to consider.

## Introduction

Suboptimal diet, characterised by factors such as low intakes of fruits and vegetables and high intakes of salt and added sugars, contributes to an estimated 90,000 deaths every year in the UK (1). More than half of over-45s are living with health conditions for which diet is a widely recognised contributing risk factor, including cardiovascular disease, type-2 diabetes, and obesity. As things stand, obesity is expected to continue increasing (2). By 2035, type 2 diabetes is projected to cost the UK National Health Services (NHS) £15 billion a year, equivalent to one and a half times as much as cancer does today (3). Halting this trajectory will be crucial for protecting the future of the UK’s health service.

Limiting intakes of salt and added sugar is one important action to prevent diet-related disease (1). Current intakes of added sugar in the UK are, on average, 55g/day (9.9 E%) in men and 44g (9.9E%) in women (4). This represents nearly double the recommendation of no more than 30g/5E% from added sugars (5). At the same time current average intakes of salt are 9.2g/ day for men and 7.6g/day for women (4), exceeding the daily recommendation of no more than 6g/day in adults (5).

Significant behaviour change would be needed to meet the recommendations for salt and added sugars in the UK. The limited success of the “5-a-day” campaign since 2003 suggests that information measures alone are not enough to change behaviour (6). In fact, research shows that upstream population-wide policies such as reformulation (as opposed to downstream interventions that depend on the individual responding) typically seem to bring about larger population-wide impacts on dietary behaviours (7,8). Supporting this research, previous modelling studies assessing the impacts of the UK’s salt reduction strategy in the early 2000s (9,10) which included reformulation, and evidence from the implementation of the UK Soft Drinks Industry Levy (SDIL) (11), suggests that reformulation measures are effective in steering the consumption of specific foods/food ingredients among consumers as well as for bringing about positive health impacts. Since the introduction of the SDIL in 2018, intake of sugar from sugar sweetened beverages (SSB) have been reduced by an average of 10%, with greater reductions amongst people on lower incomes (12). Recent poll-data shows that more than two thirds (68%) of the UK public support an extension of the SDIL to other food categories (13).

In 2021, the National Food Strategy: The Plan (NFS) (14) suggested extending the SDIL and proposed that the UK Government should introduce a £3/kg tax on sugar in foods not covered by the existing SDIL (and some ingredients used for sweetening, but not non-nutritive sweeteners) and a £6/kg tax on salt. This would impact all manufactured food categories in which sugar or salt is used as an ingredient. The report argued that this would generate an incentive for manufacturers to lower the sugar and salt content of their products, through reformulation of recipes or by reduction of portion sizes.

This study estimates the expected health benefits from the reduction in consumption of salt and sugar, and the associated reduction in energy intake, for four scenarios developed through previous work by Griffith et al (15), based on the proposed tax (14), each reflecting different manufacturer and consumer responses to a proposed extension of the SDIL to foods high in sugar and/or salt.

## Methods

### Scenarios

In this work, we adopted a methodology that combined elements of the Department of Health and Social Care (DHSC) calorie model (16) (i.e. the calculation of changes in body-mass index (BMI) across the population due to a hypothetical reduction in energy intake from sugar) with life table modelling to quantify health impacts from a reduced intake of sugar and salt in the UK. The potential sugar and salt reduction from the application of the levy proposed in the NFS has previously been calculated and described by Griffith et al. (15). Briefly, purchase panel data from the 2019 Kantar Fast Moving Consumer Goods l (Take Home) and 2016-2019 Kantar Out of Home Purchase Panel data were used to estimate changes in intakes which depend on the degrees of reformulation by the industry and the consumer response. There is considerable uncertainty about both of these aspects. Hence, four different scenarios defined by Griffith et al. (15) were tested, each reflecting different manufacturer and consumer responses to a proposed tax. Reformulation targets for different food groups set by Public Health England—i.e. a 20% reduction in sugar across all food groups (17), and food-group specific targets for salt (18)—were used to determine the four scenarios:

1. The **“Low-Low”** scenario: <u>Low</u> industry change (firms reformulate to 30% of PHE targets), and <u>low</u> consumer change (consumers substitute away from products by one third of the price increase)
2. The **“High-No”** scenario: <u>High</u> industry change (full reformulation to PHE targets), but <u>no</u> consumer change (consumers do not respond to price increases)

1. The **“High-Moderate”** scenario: <u>High</u> industry change (full reformulation to PHE targets), and <u>moderate</u> consumer change (consumers substitute away from products by 70% of the price increase)
2. The **“High-High”** scenario: <u>High</u> industry change (full reformulation to PHE targets), and <u>high</u> consumer change (consumers substitute away from products by the same amount as the price increase)

According to these previous calculations by the Institute for Fiscal Studies (14,15) a reduction in sugar intake of 4.5-12.6g per day (a 10%-29% reduction (4)) in women and 5.3-15.0g (a 10-27% reduction (4)) in men could be achieved (**Table 1**). A reduction in salt intake of 0.2-0.7g per day (a 3-9% reduction (4)) in women and 0.3-0.9g per day (a 3-10% reduction(4)) in men could be achieved by introducing the levy. More details about how the scenarios were defined can be found in published reports (14,15).

**Table 1.** Scenarios tested, based on previous work(15).

|  |  | **Salt**<br>**reduction**<br>**(g)** | **SD** | **Sugar**<br>**reduction**<br>**(g)** | **SD** | **Kcal**<br>**reduction** | **SD** | **Weight**<br>**reduction<sup>a</sup>**<br>**(kg)** | **SD** |
| --- | --- | --- | --- | --- | --- | --- | --- | --- | --- |
| Low-Low | Males+18 | -0.3 | 0.2 | -5.3 | 3.6 | -21.2 | 14.4 | -0.9 | 0.6 |
|  | Females+18 | -0.2 | 0.1 | -4.5 | 3.2 | -18.0 | 12.8 | -0.8 | 0.5 |
| High-No | Males+18 | -0.7 | 0.5 | -9.7 | 7.0 | -38.8 | 28 | -1.6 | 1.2 |
|  | Females+18 | -0.6 | 0.5 | -8.3 | 6.0 | -33.2 | 24 | -1.4 | 1.0 |
| High-Moderate | Males+18 | -0.8 | 0.6 | -13.6 | 9.2 | -54.4 | 36.8 | -2.3 | 1.5 |
|  | Females+18 | -0.7 | 0.4 | -11.4 | 8.0 | -45.6 | 32 | -1.9 | 1.3 |
| High-High | Males+18 | -0.9 | 0.6 | -15.0 | 10.2 | -60.0 | 40.8 | -2.5 | 1.7 |
|  | Females+18 | -0.7 | 0.5 | -12.6 | 8.7 | -50.4 | 34.8 | -2.1 | 1.5 |

<sup>a</sup>Assuming that 0.042 kg body weight is lost per calorie reduction (22).

Based on previous evidence, changes in salt consumption were hypothesised to impact health outcomes directly (stomach cancer, (19)) and indirectly (ischemic heart disease (IHD) and stroke via hypertension (20)) (Figure 1). Changes in sugar consumption were hypothesised to impact health outcomes both directly (IHD,(19,21)), and indirectly (through BMI (21)). Hence, like the DHSC model, our model also considered health impacts from changes in BMI resulting from a decreased intake of calories from sugar (Figure 1).

**Figure 1.**
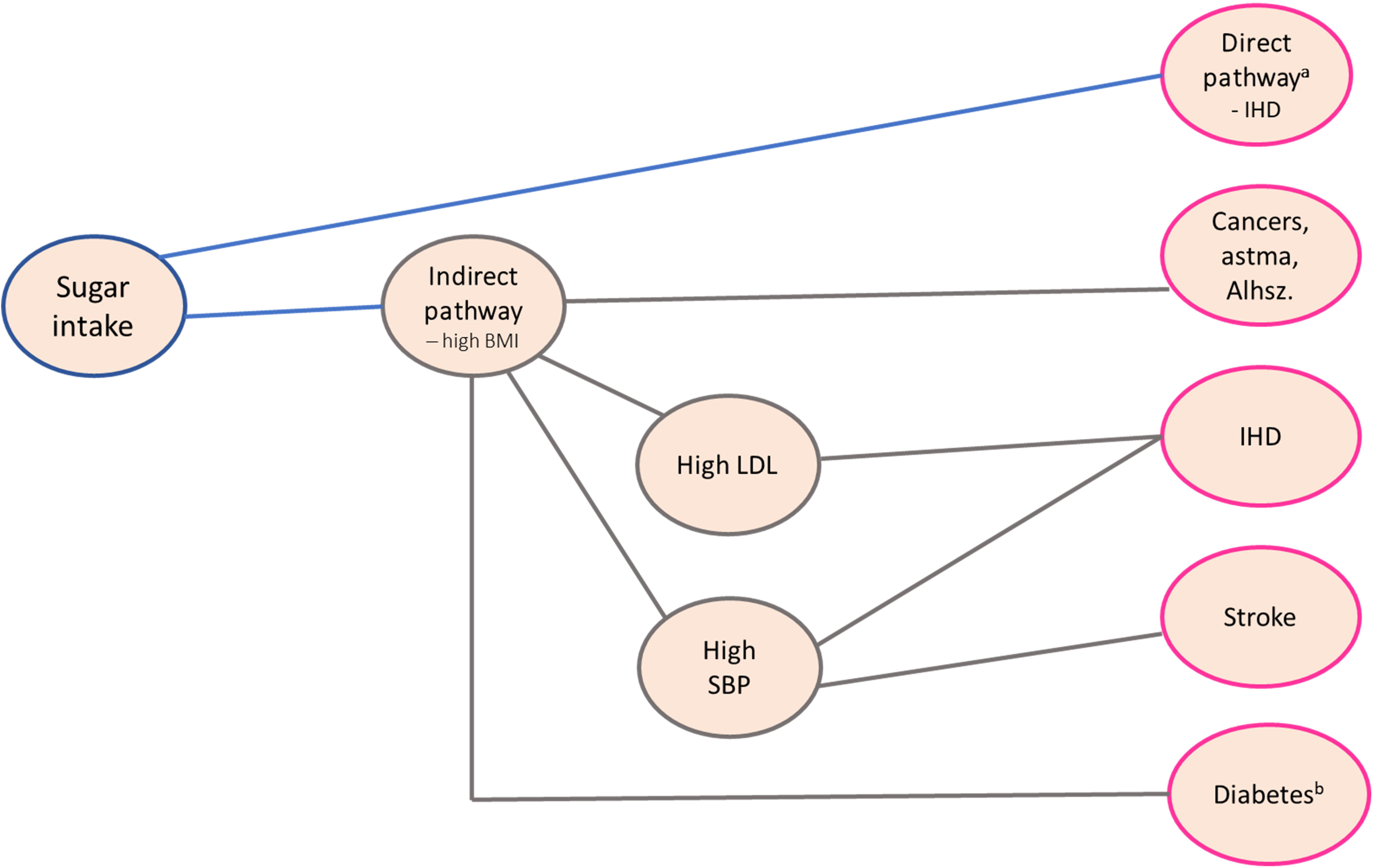
Pathways through which sugar and salt intake affects disease outcomes directly and indirectly. ^a^Unregulated hepatic fructose metabolism leading to hepatic de novo lipogenesis which in turn promotes very low-density lipoprotein production (21). ^b^High fasting glucose levels from insulin resistance due to high free fatty acid levels (and the predominant utilisation of lipids by muscle and skeleton) leading to hyperinsulinemia and ultimately also impaired insulin secretion (21). BMI, body-mass index; SBP, systolic blood pressure; LDL, low density lipoprotein; IHD, ischemic heart disease.

All the scenarios used the following input parameters for the health impact modelling:

- Policy lifetime and evaluation period = 25 years. The levy is implemented immediately and is active for 25 years, thus the health-related impacts of the levy are also evaluated over 25 years. This timeline was chosen to mirror that used for the NFS (14), but also because this is long enough to show significant health impacts from the policies in question. However, impacts are likely to continue beyond 25 years, so the net present value of these policies could increase further as more years are added to the model.
- The health impacts are quantified for the entire adult (18+) population in the UK.
- Scenarios have age and gender specific calorie reductions, based on previous work by the Griffith et al. (15) and given in Table 2.

**Table 2.** Disease outcomes used for the modelling.

| **Disease outcomes** | **Dietary**<br>**risk** | **Relative risk** | **Change in**<br>**units** | **Dose-response**<br>**relationship** |
| --- | --- | --- | --- | --- |
| Ischemic heart disease<sup>a</sup> | Salt | 0.510 | -6 | log-linear |
| Ischemic heart disease<sup>b</sup> | Sugar | 0.982 to 0.941 | -4.9 to -13.80 | non-linear |
| Ischemic heart disease | BMI | 0.587 | -5 | log-linear |
| Ischemic stroke | Salt | 0.390 | -6 | log-linear |
| Ischemic stroke | BMI | 0.556 | -5 | log-linear |
| Stomach cancer | Salt | 0.976 to 0.924 | -0.25 to -0.80 | non-linear |
| Oesophageal cancer | BMI | 0.730 | -5 | log-linear |
| Colorectal cancer | BMI | 0.898 | -5 | log-linear |
| Type 2 diabetes | BMI | 0.368 | -5 | log-linear |
| Gallbladder cancer | BMI | 0.804 | -5 | log-linear |
| Pancreatic cancer | BMI | 0.925 | -5 | log-linear |
| Breast cancer | BMI | 1.124 | -5 | log-linear |
| Uterine cancer | BMI | 0.628 | -5 | log-linear |
| Ovarian cancer | BMI | 0.976 | -5 | log-linear |
| Kidney cancer | BMI | 0.782 | -5 | log-linear |
| Thyroid cancer | BMI | 0.850 | -5 | log-linear |
| Multiple Myeloma | BMI | 0.917 | -5 | log-linear |
| Acute lymphoid leukaemia | BMI | 0.902 | -5 | log-linear |
| Chronic lymphoid leukaemia | BMI | 0.902 | -5 | log-linear |
| Chronic myeloid leukaemia | BMI | 0.902 | -5 | log-linear |
| Other leukaemia | BMI | 0.902 | -5 | log-linear |
| Intracerebral haemorrhage | BMI | 0.470 | -5 | log-linear |
| Subarachnoid haemorrhage | BMI | 0.470 | -5 | log-linear |
| Hypertensive heart disease | BMI | 0.404 | -5 | log-linear |
| Atrial fibrillation and flutter | BMI | 0.743 | -5 | log-linear |
| Asthma | BMI | 0.712 | -5 | log-linear |
| Alzheimer | BMI | 0.822 | -5 | log-linear |

<sup>a</sup>RRs for ischemic heart disease (IHD) and stroke were derived from previous research (20).
<sup>b</sup>RRs for sugar sweetened beverages (SSB) from GBD (19) were used as a basis for the sugar-IHD relationship. Data from previous research (55) was used to estimate the average sugar content/100 ml of the SSB consumed in the UK. The average sugar content/100 ml of High-sugar, Mid sugar and Low-sugar beverages was used to compute a weighted average sugar content/100 ml of SSB. This weighted average (=7.86 g/100ml) was used as a proxy for sugar intake and was thus applied to model the reduction of sugar.

The primary outcomes of the modelling were: i) changes in life expectancy and Years of Life Gained (YLG) from changes in mortality accumulated over 25 years and changes in morbidity (new cases of different diseases) accumulated over 25 years.

### Quantifying reductions in kcal and BMI

In accordance with the DHSC calorie model, 1 gram of reduced sugar intake per day was translated to 4 kcal reduced energy intake from sugar. Each kcal reduction was in turn assumed to correspond to a 0.042kg reduction in body weight using the steady-state models originally developed by Kevin Hall *et al* (22).

The UK National Diet and Nutrition Survey 2019 (4) was used to provide a nationally representative adult cohort (N=586) to which changes in body weight (and BMI) were applied. Each adult in the survey cohort was first assigned their (baseline) BMI based on their height and weight. Similar to the DHSC approach, new body weights (and thus new BMIs) resulting from a reduced energy intake from sugar were calculated. New bodyweights were calculated for adults considered overweight (defined as BMI ≥ 25 kg/m2) only. This is because we assumed that reductions in calorie intake would have minimal impact on the weight of healthy weight and underweight people as they are thought to generally be highly effective at maintaining weight stability, even with fluctuations in calorie intake (23). The average BMI of the entire baseline adult population was then compared to the average BMI of the adult population after modifications to weights/BMIs were computed under each of the four scenarios. The difference between the average BMI of the total adult population at baseline and the average BMI of the total adult population when the BMI of overweight/obese adults was changed were used as inputs in the life table model.

### Disease outcomes and relative risks

Dose-response relationships (i.e. relative risks per unit change in consumption) between dietary intake and chronic disease morbidity and mortality were obtained from the 2019 Global Burden of Disease (GBD) study (19). The relative risk (RR) for a given dietary risk-disease pair estimates how much the risk of mortality (or morbidity) would change when the dietary risk changes. For example, the risk of IHD is reduced by 49% for each 6 g reduction in salt intake (Table 2). A total of 24 disease outcomes (Table 2) were considered for the modelling as they have been shown to be linked to sugar/salt consumption and/or BMI (19). Further details regarding the relative risks may be found in the Supplementary Methods.

### Health impact modelling

Health impacts were the main outcome of this study, and calculated on the basis of a previously applied methodology (24), using the life table model IOMLIFET (25), implemented in R (26). Life tables were separately generated for males and females, due to their different underlying mortality and morbidity rates. The latest (2019) data on age-specific and sex-specific population-size estimates, and on age-specific and sex-specific all-cause mortality were obtained from the Office for National Statistics (27). Data on disease-specific mortality (mortality rates) and morbidity (incidence rates) for relevant outcomes (Table 3) were downloaded from the GBD results tool (28). Together, these data provided the baseline input data for the UK.

**Table 3.** Scenario specific reductions in calories, corresponding reductions in body weight, baseline BMI, average adult BMI resulting from a reduced energy intake from sugar and difference between baseline vs. modelled BMI, grouped by gender.

| Average<br>BMI<br>baseline | BMI<br>change | Uncertainty<br>range<sup>a</sup> |
| --- | --- | --- |
| 27.61 | -0.20 | -0.06 to -0.34 |
| 27.15 | -0.16 | -0.04 to -0.27 |
| 27.61 | -0.37 | -0.1 to -0.64 |
| 27.15 | -0.29 | -0.08 to -0.51 |
| 27.61 | -0.52 | -0.17 to -0.87 |
| 27.15 | -0.40 | -0.12 to -0.69 |
| 27.61 | -0.57 | -0.18 to -0.97 |
| 27.15 | -0.44 | -0.14 to -0.75 |

<sup>a</sup>Range represents lower and upper estimates for changes in BMI based on a dispersion measure of +/-1SD around the mean for reductions in sugar consumption among males and females, respectively, combined with upper and lower 95% confidence intervals for relevant RRs provided by the Global Burden of Disease study (28).

Life table models were used to quantify changes in life expectancy and Years of Life Lost (YLL) from changes in dietary risk exposures according to the four different reformulation scenarios. Briefly, the IOMLIFET model estimates survival patterns in the population over time based on age-specific mortality rates. Based on the information of a hypothetical change in diet (risk-exposure) and a known exposure-response function (i.e. relative risk), changes in survival rates can be quantified as e.g., YLL or changes to life expectancy. YLL can be explained as the years of life lost for an individual (or a population) as a result of premature avertable mortality, considering the age at which deaths occurred. Since the dietary modifications were expected to reduce mortality rates, YLL were translated to years of life gained (YLG).

Changes in salt and sugar consumption were assumed to be adopted instantly while underlying mortality and incidence rates remained constant for the duration of follow-up. Our modelling strategy also did not allow for general increases in BMI in the population over time, and we assumed the same number of new live births into the future. The effects on ischemic heart disease, stroke, and type 2 diabetes were assumed to reach their maximum impact after 10 years, while the maximum impact for cancers was expected after 30 years, with no change in cancer risk during the first 10 years. To account for the time delays between dietary changes and health outcomes, we used time-varying functions based on cumulative distribution curves. In cases where several dietary exposures affected the same disease, the risks were multiplied together as done previously (24). Changes in YLG were quantified as the additional number of years of life that individuals in the UK adult population would live as a result of reducing their risk of dying prematurely from disease outcomes relating to salt and sugar consumption. Changes in morbidity (new cases of disease) were quantified using the same principles. Morbidity calculations were performed using the output population from the life table as the baseline population, to which changes in risk exposures were applied. Each morbidity calculation was performed separately based on incidence rates and summed over the UK adult population. Changes in life expectancy at birth were calculated as the difference between the baseline life expectancy (the expected life years divided by the starting population) and the impacted (modelled) life expectancy (the impacted expected life years divided by the impacted starting population).

### Sensitivity analysis

To test the sensitivity of the results to key parameters, we generated upper and lower health impact estimates based on a dispersion measure of +/-1SD around the mean for reductions in salt and sugar (later translated to changes in BMI), (Table 1), combined with upper and lower 95% CIs for the RRs provided by the GBD (Supplementary Table 1). More information regarding data management and assumptions for the health impact modelling may be found in the Supplementary Methods.

## Results

### Changes in energy intake and BMI

Based on the four scenarios, and described assumption, the energy intake from sugar was translated to a change in body weight ranging from −0.8 to −2.1kg in men, and −0.9 to −2.5kg in women (Table 1) with half of the weight change being achieved in about 1 year and 95% of the weight change in about 3 years (22). BMI changes ranged from −0.17 to −0.48 amongst men and −0.19 to −0.53 amongst women (Table 3). Changes in the distribution of BMI across the adult population in the UK after the reduced consumption in sugar are displayed in Supplementary Figure 1.

In the High-High scenario, the prevalence of overweight and obesity dropped from 69 to 62 percent and from 54 to 49 percent from among males and females, respectively (no data shown).

### Health impacts

The results of the health impact modelling show that average life expectancy in the UK could increase by up to 4.9 (1.0-9.4) months if there was both a high manufacturer and high consumer response to the proposed tax on salt and sugar (Table 4). Even with a minimal response from both manufacturers and consumers, the estimated gain in life expectancy would be 1.7 (0.3-3.6) months. Total YLG accumulated over 25 years from avoided deaths ranged from 1.2 (0.3-2.5) million in the Low-Low scenario to 3.5 (0.8-6.4) million in the High-High scenario, with similar numbers of QALYs being accumulated through reduced morbidity. The largest reductions in new cases of disease came from cardiovascular disease (up to 1.0 (0.2-1.9) million fewer cases), followed by type 2 diabetes, with smaller reductions in cancers and respiratory disease (Table 4).

**Table 4.** Results from life table modelling based on mortality and morbidity, combining health.

| Outputs | Low-Low | High-No | High-Moderate | High-High |
| --- | --- | --- | --- | --- |
|  | Central estimate<br>(uncertainty range<sup>d</sup>) | Central estimate<br>(uncertainty range<sup>d</sup>) | Central estimate<br>(uncertainty range<sup>d</sup>) | Central estimate<br>(uncertainty range<sup>d</sup>) |
| Changes in LE in months | 1.7<br>(0.3-3.6) | 3.6<br>(0.6-7.2) | 4.5<br>(0.9-8.8) | 4.9<br>(1.0-9.4) |
| Total YLG in millions<sup>a</sup> | 1.2<br>(0.3-2.5) | 2.6<br>(0.5-5.1) | 3.2<br>(0.7-6.0) | 3.5<br>(0.8-6.4) |
| Reduction in cases of disease<sup>b</sup> |  |  |  |  |
| Cardiovascular disease<sup>c</sup> | -348 887<br>(-75 605 to -728 811) | -742 739<br>(-134 718 to -1 474 625) | -942 611<br>(-196 088 to -1 778 274) | -1 012 056<br>(-227 208 to -1 901 444) |
| Diabetes | -208 093<br>(-44 281 to -427 517) | -374 105<br>(-78 950 to -773 318) | -523 513<br>(-127 320 to -1 033 524) | -571 310<br>(-139 858 to -1 123 479) |
| Cancers | -4 271<br>(-732 to -12 468) | -6 837<br>(-1 157 to -25 422) | -10 459<br>(-1 568 to -37 304) | -11 812<br>(-1 864 to -41 360) |
| Respiratory disease | -90 605<br>(-18 043 to -195 572) | -165 521<br>(-33 291 to -355 432) | -230 354<br>(-53 183 to -492 122) | -249 126<br>(-59 122 to -538 795) |

impacts from both reduced salt and sugar consumption.
<sup>a</sup>Based on changes in mortality, accumulated over 25 years.552 <sup>b</sup>Changes in incidence of disease, accumulated over 25 years.553 <sup>c</sup>IHD, ischemic stroke, intracerebral/subarachnoid haemorrhage, hypertensive heart disease, atrial fibrillation and flutter.554 <sup>d</sup>Range represents upper and lower health impact estimates based on a dispersion measure of +/-1SD around the mean for reductions in salt and sugar consumption among males and females, respectively, combined with upper and lower 95% confidence intervals for relevant RRs provided by the Global Burden of Disease study (28).
LE = Life expectancy.
YLG = Years of Life GainedQALY = Quality Adjusted Life Year.

Differences in YLG between the four scenarios show that the High-Moderate and High-High scenarios are relatively similar in terms of mortality impacts, and that even the scenario assuming no consumer response to the taxes would achieve around 75% of the benefit of the High-High scenario that assumes a high degree of consumer response (Figure 2). The Low-Low scenario assuming a low degree of manufacturer as well as consumer response would achieve less than half of the benefits of the other scenarios.

**Figure 2.**
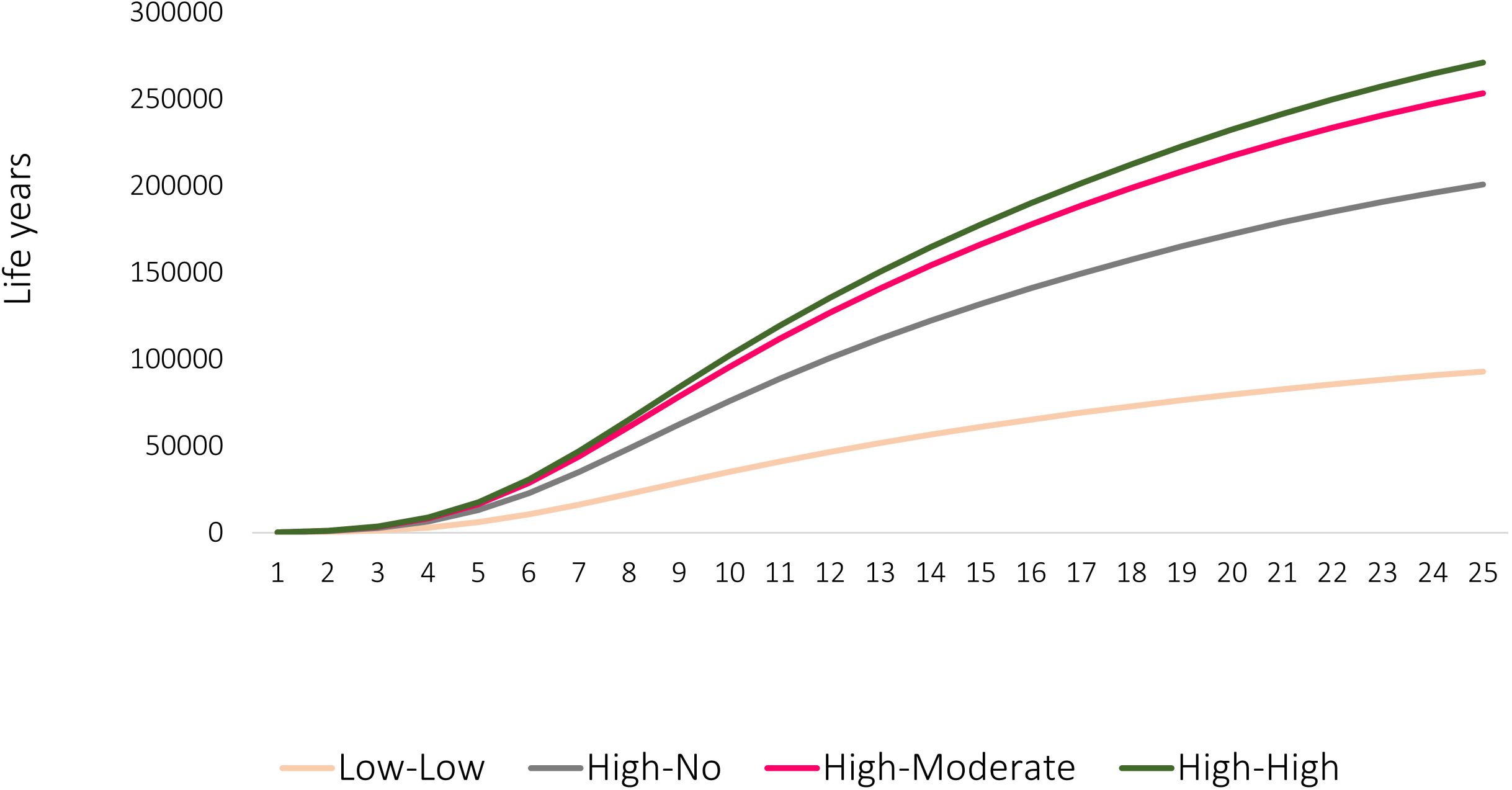
Years of life gained each year up to year 25 for the four modelled scenarios.

Slightly over half of the gains in YLG (48-54% depending on the scenario) can be attributed to the reduction in sugar consumption (both direct and indirect pathways) and more than half of those impacts can be ascribed to a reduced BMI in the UK population (Figure 3) with the remainder accruing from direct impacts of sugar consumption on YLG.

**Figure 3.**
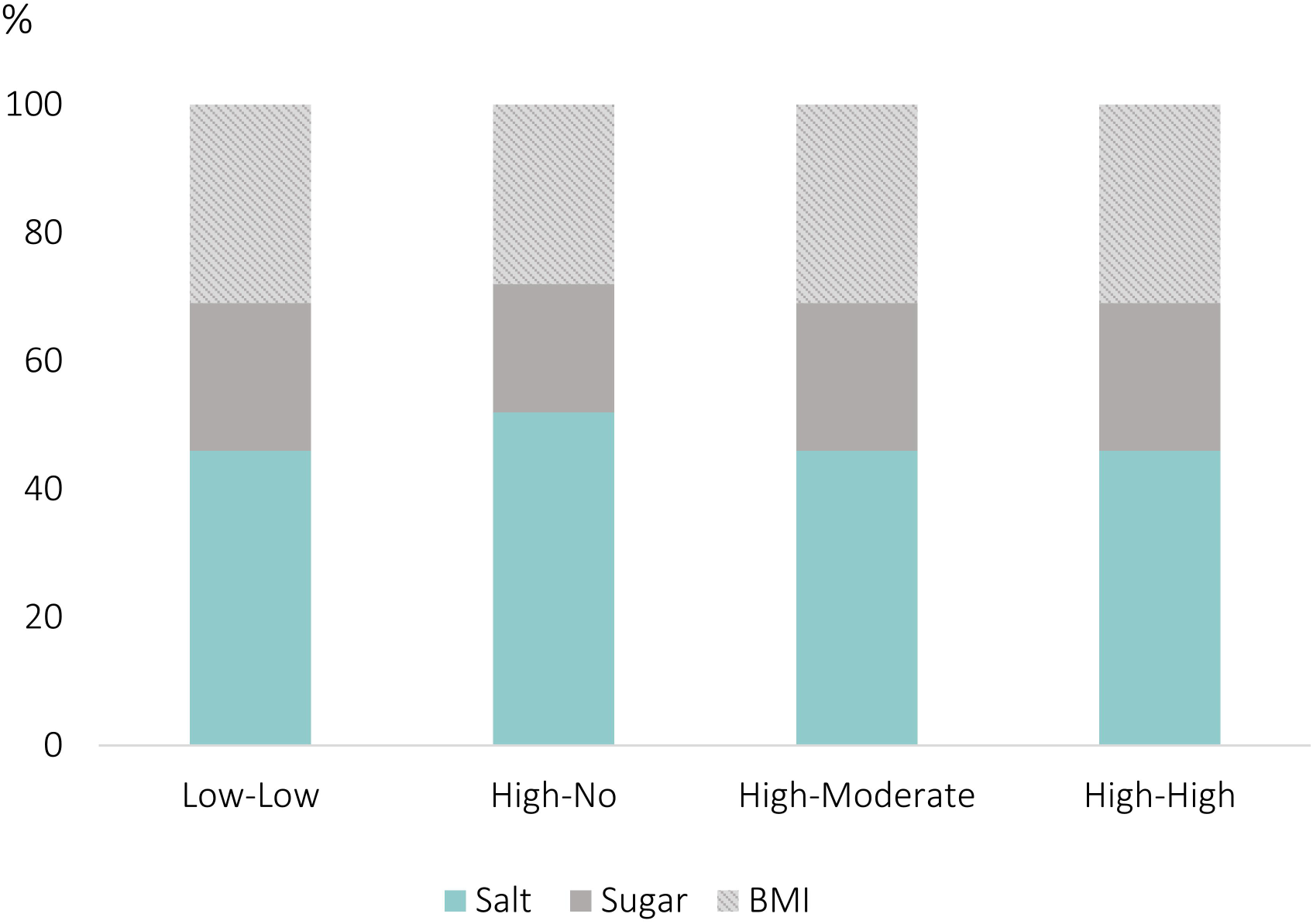
Share (%) of years of life gained attributed to each risk factor.

## Discussion

The results of this modelling show that extending the current soft drinks levy to other foods high in sugar and adding a tax on salt could lead to significant health benefits for the UK. Life expectancy in the UK could be increased by between 1.7 months and nearly 5 months, depending on the degree of industry and consumer response to the tax. In the best-case scenario, with a high response from both consumers and industry, the total number of preventable cases of chronic disease could be reduced almost 2 million over 25 years, including reductions in CVD, type 2 diabetes, respiratory disease and cancer. The main part of these gains can be attributed to reduced mortality and morbidity from cardiovascular diseases, which remain the commonest causes of death and disability in the UK and are strongly associated with consumption of salt and sugar.

Our results show that a 29 and 27 percent reduction in sugar intake resulting from the tax among males and females, respectively, could reduce overweight and obesity by as much as 10.9 and 10.5 percent in males and females, respectively. This is similar to other research where the prevalence of obesity was reported to reduce by 1.5 to 10 percent when added sugar intake reduced by 10 to 20 percent (29,30). Our results are also consistent with previous modelling studies showing improved health outcomes from diets that better align with dietary recommendations (31,32), and tally with the findings of previous modelling studies that have shown health gains resulting from a reduced intake of added sugars specifically (33,34). Health gains from modelling studies have also been reported as a result of the taxation of salt or foods high in salt in other countries (35,36). For example, taxation of sodium in the United States has been estimated to potentially reduce sodium intake by 9.5% in the population, and could avert nearly a million cardiovascular events over the lifetime of adults aged 40 to 85, increasing QALYs by 2.1 million (37). Our findings add to this previous body of research, and thus reinforce arguments for why currently high intakes of salt and sugar should be targeted using fiscal policies.

The UK SDIL has already demonstrated its impact on reducing sugar intake at a national level (12). Our findings further support global trends and recommendations regarding fiscal policies aimed at the taxation of foods. A recent WHO publication (38), reviewing the existent evidence on the taxation of foods, (conditionally) recommends the implementation policies to tax foods that do not contribute to a healthy diet. The recommendation is based on findings from countries such as Mexico (39), Hungary (40), and Denmark (41), which have all suggested decreased purchases of taxed foods from food categories such as confectionery and ice cream. These international examples reinforce the potential impact of an expanded taxation strategy in the UK as a means to curb excessive salt and sugar intake and enhance overall public health. However, simply implementing food taxes may not be the most effective strategy, as multicomponent approaches that incorporate a mix of both upstream and downstream interventions, typically generate the greatest reductions in population-wide salt and sugar consumption (7,8). For example, the UK Food Standards Agency’s campaign (2003–2011) on salt reduction, involving a combination of awareness campaigns, agreed target settings, voluntary reformulation from industry and population monitoring of salt consumption, led to a 1.4g per day reduction in population salt intake in England (42). This reduction is significantly larger than the one estimated from the salt levy proposed in the NFS (15), which was incorporated into our modelling.

To the best of our knowledge, this is the first study to explore the health effects of the salt and sugar tax proposed by the NFS. This analysis brings together different methods and data sources to explore co-benefits of various pathways of reduced salt and sugar consumption. An important strength is the ability to assess both direct and indirect pathways through which these consumption changes would lead to changes across time and dimensions of health and cost. Compared to looking at indirect vs. direct pathways in isolation, these analyses are therefore likely to capture a more complete picture of the possible health benefits of fiscal measures that reduce intakes of salt and of added sugars. UK dietary recommendations (the Eatwell Guide) recommend that adults limit intakes of salt and added sugars to no more than 6g/d and 30g/d, respectively (5). Given the UK current average consumption being considerably higher than that, our modelled scenarios, leading to reductions of up to 10% in salt and 29% in sugar, while ambitious, are metabolically rational and concordant with governmental goals.

A limitation to our modelling is that we made no assumptions about possible dietary substitution in our analyses. The estimates of salt and sugar reductions by Griffith et al. (15) are not based on price elasticities or demand models and do, therefore, not take account of complex substitution behaviour. Removal of salt and added sugars in the diet could result in subsequent substitution with other foods that are instead high in e.g. other sweeteners or saturated fats. Unfortunately, there is currently no up-to date published data on substitution behaviour based on a demand model or estimates of price elasticities. The calculations on salt and sugar reductions by Griffith et al. (15) only assume different levels of the overall elasticity of sugar purchased to the tax. To date, evidence from intervention and modelling studies concerning what happens to consumption of other foods if intakes of salty and sugary foods reduce is sparse. To our knowledge, the only estimates based on a demand model that provide those are in Tiffin et al. (43), and Harding and Lovenheim (44). However, the former are from 2011 and thus out of date, and the latter are based on the US context and are not representative for UK consumers. Recent analyses of food consumption trends in the UK show that intakes of sugar have declined while intakes of fruits, and vegetables in particular, have increased over the last 40 years (45). Furthermore, several studies indicate that fruit consumption can replace consumption of sweet snacks (46). Therefore, our models are more likely to have underestimated the health impacts of the proposed tax due to substitution with healthy foods as opposed to overestimating them due to substitution with other unhealthy foods. Measures such as subsidies to reduce the cost of fruits and vegetables could be combined with a salt and sugar tax to facilitate and encourage health-promoting substitutions. In fact, previous studies that explored the combined effects of taxes for unhealthy foods/food components with subsidies (vs. taxes only) reported larger health benefits (35,47). Combining taxes with subsidies would also help avoid constraining the resources of the lowest-income households that already dedicate >15% of total budgets to food purchases (48). The funds raised by the tax could be spent on complementary measures addressing current inequalities around food, such as e.g., an expansion of the free school meal scheme, public health initiatives, and subsidisation of, and increasing access to, health promoting foods.

Despite being adjusted for age, sex, smoking, and physical activity, using RRs in our modelling of health impacts is a limitation because of the possibility for residual confounding. In our case, we applied 95% CIs for the RRs to provide a measure of variation in the selected health outcomes, but these may overestimate the CI widths due to aggregation across disease endpoints. There are also uncertainties related to the survey data used to develop the scenarios. These data are based on purchases and do thus not necessarily accurately capture people’s actual consumption. Moreover, we only modelled the adult population. Considering the rise in childhood obesity in the UK (49), it is likely that the inclusion of children with type-2 diabetes would have generated additional health benefits. Our model also does not take into consideration the effects of socioeconomic status or ethnic origin. Because dietary patterns and disease burdens vary across sociodemographic/economic groups in the UK (50), this could have influenced our final outputs. Lastly, the wide uncertainty intervals of our results indicate considerable variability in the estimates, which may affect the robustness and generalizability of our findings. However, they could help estimate the likely extent of health benefits from proposed interventions, providing essential information for health policymakers as they evaluate various options for reducing salt and sugar intake in the UK.

This study highlights the significant health benefits that could be achieved by a fiscal measure that reduces consumption of salt and added sugars in UK adults. Despite the potential benefits, food taxes are complex and sometimes controversial, and it is important to consider possible barriers to implementation/effective implementation of any proposed tax. For example, a sugar tax may lead manufacturers to substitute fat for sugar which may have negative consequences for health. Furthermore, industry acceptability is likely to be crucial for avoiding increased costs to consumers, especially to low-income families. Combining taxes with subsidies could be a way to help avoid business losses and thus increase industry acceptability (51). Equally important is consumer acceptability. Previous research shows that taxes are generally unpopular and seen as intrusive (52,53). However, it is promising that substantial public support seems to exist in the UK for an extension of the SDIL to other food categories (13). Lastly, the success of the proposed tax is also likely to depend on the degree of the management, accountability and public resources available to enable its implementation (54).

By investigating the effects of a combined salt and sugar tax applied to a wide range of food categories, and through a range of different plausible scenarios of manufacturer and consumer response, our work adds new evidence to support the introduction of targeted fiscal measures to reduce both salt and sugar consumption in the UK. These benefit estimates are not definitive and carry significant uncertainty. Nevertheless, they should help determine the order of magnitude of health benefits that might be expected, and thus lend important weight to support health policy makers in their review of options to implement salt and sugar reduction.

## Data Availability

All data produced in the present study are available upon reasonable request to the authors.

## Abbreviations

AF: Atrial Fibrillation
BMI: Body mass index
DHSC: Department of Health and Social Care
GBD: Global Burden of Disease
IHD: Ischemic heart disease
NFS: National Food Strategy
NHS: National Health Services
RR: Relative Risk
SDIL: Sugary drinks industry levy
SSB: Sugar sweetened beverages
YLG: Years of Life Gained
YLL: Years of Life Lost

## Notes

### Competing Interest Statement

The authors have declared no competing interest.

### Funding Statement

This project received funding from Impact on Urban Health. PEC had financial support from the Swedish Research Council (VR, grant nr. 2022-00344) for the submitted work. The funders had no role in the design of the study; in the collection, analyses, or interpretation of data; in the writing of the manuscript, or in the decision to publish the results.

### Summary of Updates

A few new references added to the introduction and discussion, and a new supplementary table (#2) has been included.

