## Supplementary information for "Population health impacts from the taxation of salt and sugar in the United Kingdom"

### Materials and methods

#### **Supplementary Figure 1.** Distribution of body-mass index in the adult UK population at baseline and after weight-loss resulting from reduced sugar intake in the High-High scenario.

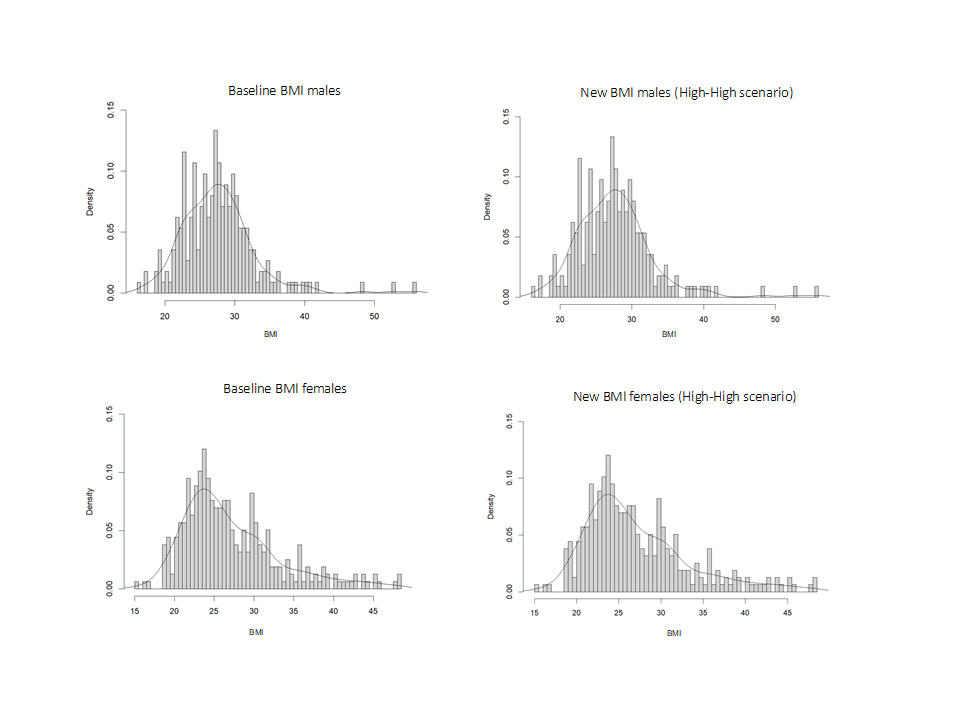

#### Handling input data and relative risks

The disease-specific mortality and morbidity data were only available by 5-year age ranges and not in single-year-of-age format. Using the weighted mean age of the UK population in each interval, the rate for each 1-year age interval was interpolated using monotonic cubic spline interpolation using the programme SRS splines, available as an add-in to Excel [1].

Relative risks from the GBD which were expressed in terms of a harmful risk factor (e.g. “diet high in sodium/salt”) were inverted to create relative risks for a positive change in diet. The GBD provides relative risks for females and males combined, by 5-year age ranges, starting from the age of 25. The relative risks for each risk, outcome, and 5-year age range were weighted according to the age distribution in the UK population to create one single relative risk for each risk-outcome pair. Ages from 18-25 were assigned the same relative risk as the 25-29 age group, although the baseline mortality from the included dietary risks was almost non-existent in the UK population aged 18-25. The RRs for IHD-sugar and stomach cancer-salt were non-linear, i.e. different RRs were provided for different, pre-defined levels of risk. Therefore, monotonic cubic spline interpolation [1] was applied to obtain the RR reflecting the specific changes in salt and sugar intake as per the four scenarios.

#### Time lags implemented in the modelling

Previous research assessing effects of dietary interventions on various causes of mortality has established approximate time lags between exposure and onset of disease [2, 3]. Hence, effects on ischemic heart disease, stroke and type 2 diabetes were assumed to reach a maximum impact after 10 years and for cancers after 30 years, with no change in cancer risk during the first 10 years. Time-varying functions based on cumulative distribution functions of normally distributed variables (s-shaped curves) were implemented to account for time lags between dietary changes and changes in health outcomes. Details with regards to the implementation of the time-lags have previously been described [4].

#### **Supplementary Table 1.** Disease outcomes and associated relative risk (lower and upper confidence intervals) used for the modelling.

| **Disease outcomes** | **Dietary risk** | **Relative risk**  **(lower CI-higher CI)** | **Change in units** | **Dose-response relationship** |
| --- | --- | --- | --- | --- |
| Ischemic heart disease^a^ | Salt | 0.500-0.530 | -6 | log-linear |
| Ischemic heart disease^b^ | Sugar | 0.968 to 0.914-1.000 to 1.000 | -4.9 to -13.80 | non-linear |
| Ischemic heart disease | BMI | 0.487-0.776 | -5 | log-linear |
| Ischemic stroke | Salt | 0.360-0.410 | -6 | log-linear |
| Ischemic stroke | BMI | 0.460-0.718 | -5 | log-linear |
| Stomach cancer | Salt | 0.971 to 0.907-0.997 to 0.990 | -0.25 to -0.80 | non-linear |
| Oesophageal cancer | BMI | 0.572-0.959 | -5 | log-linear |
| Colorectal cancer | BMI | 0.876-0.992 | -5 | log-linear |
| Type 2 diabetes | BMI | 0.294-0.476 | -5 | log-linear |
| Gallbladder cancer | BMI | 0.728-0.892 | -5 | log-linear |
| Pancreatic cancer | BMI | 0.871-0.982 | -5 | log-linear |
| Breast cancer | BMI | 1.094-1.151 | -5 | log-linear |
| Uterine cancer | BMI | 0.602-0.656 | -5 | log-linear |
| Ovarian cancer | BMI | 0.940-1.014 | -5 | log-linear |
| Kidney cancer | BMI | 0.739-0.825 | -5 | log-linear |
| Thyroid cancer | BMI | 0.787-0.925 | -5 | log-linear |
| Multiple Myeloma | BMI | 0.866-0.970 | -5 | log-linear |
| Acute lymphoid leukaemia | BMI | 0.860-0.946 | -5 | log-linear |
| Chronic lymphoid leukaemia | BMI | 0.860-0.946 | -5 | log-linear |
| Chronic myeloid leukaemia | BMI | 0.860-0.946 | -5 | log-linear |
| Other leukaemia | BMI | 0.860-0.946 | -5 | log-linear |
| Intracerebral haemorrhage | BMI | 0.365-0.629 | -5 | log-linear |
| Subarachnoid haemorrhage | BMI | 0.365-0.629 | -5 | log-linear |
| Hypertensive heart disease | BMI | 0.266-0.655 | -5 | log-linear |
| Atrial fibrillation and flutter | BMI | 0.678-0.816 | -5 | log-linear |
| Asthma | BMI | 0.650-0.760 | -5 | log-linear |
| Alzheimer | BMI | 0.711-0.952 | -5 | log-linear |

^a^RRs for ischemic heart disease (IHD) and stroke were derived from previous research [5].

^b^RRs for sugar sweetened beverages (SSB) from GBD [6] were used as a basis for the sugar-IHD relationship. Data from previous research [7] was used to estimate the average sugar content/100 ml of the SSB consumed in the UK. The average sugar content/100 ml of High-sugar, Mid sugar and Low-sugar beverages was used to compute a weighted average sugar content/100 ml of SSB. This weighted average (=7.86 g/100ml) was used as a proxy for sugar intake and was thus applied to model the reduction of sugar.

#### **Supplementary Table 2.** Main assumptions of the health impact model.

|  | **Main assumptions** | **Place in text** |
| --- | --- | --- |
| 1 | Relative risks for each risk, outcome, and 5-year age range were weighted according to the age distribution in the UK population | Page 3 Supplementary Materials |
| 2 | Relative risks from the GBD were inverted to create relative risks for a positive change in diet | Page 3 Supplementary Materials |
| 3 | Disease-specific mortality and morbidity data was interpolated using monotonic cubic spline interpolation to get the rate for each 1-year age interval | Page 3 Supplementary Materials |
| 4 | Changes in salt and sugar consumption were assumed to be adopted instantly | Page 9 main text |
| 5 | Underlying mortality and incidence rates remained constant for the duration of follow-up | Page 9 main text |
| 6 | General increases in BMI in the population over time were not allowed | Page 9 main text |
| 7 | The same number of new live births into the future were assumed | Page 9 main text |
| 8 | The effects on ischemic heart disease, stroke, and type 2 diabetes were assumed to reach their maximum impact after 10 years, while the maximum impact for cancers was expected after 30 years, with no change in cancer risk during the first 10 years | Page 9 main text and page 3 Supplementary Materials |
| 9 | To account for the time delays between dietary changes and health outcomes, time-varying functions based on cumulative distribution curves were used | Page 9 main text and page 3 Supplementary Materials |
| 10 | In cases where several dietary exposures affected the same disease, the risks were multiplied together | Page 9 main text |
